# Are Frontier Large Language Models Safer Than Government-Backed Symptom Checkers for Clinical Self-Triage? A Standardised Vignette Evaluation

**DOI:** 10.64898/2026.09.01.26361908

**Authors:** Azwad Raza Chowdhury, Bushra Chowdhury

## Abstract

**Background:** Consumer use of AI chatbots for health advice is rising, yet triage safety relative to established services remains unclear. Australia’s Healthdirect, a government-backed symptom checker with 2.4 million uses in FY2024–25, remains unevaluated against frontier large language models (LLMs), and whether premium subscriptions improve triage safety remains unexplored. This study compared the triage accuracy and safety of Healthdirect against six LLM configurations across ChatGPT, Claude, and Gemini, assessed whether paid subscriptions improve triage safety, and characterised each system’s error patterns.

**Methods:** Forty-five clinical vignettes from the Semigran et al. benchmark spanning emergency, non-emergent, and self-care categories (15 each) were evaluated across seven systems. Healthdirect was tested following a seven-rule interaction protocol. LLMs were evaluated using first-person patient-language prompts under free-tier and paid-tier conditions. Outcomes were triage accuracy, emergency sensitivity, under-triage, and critical misses, analysed using Cochran’s Q, Bonferroni-corrected McNemar tests, Cohen’s kappa, and Wilson intervals.

**Findings:** Triage accuracy differed significantly (Cochran’s Q = 36.79, p < 0.001). Healthdirect achieved 48.9% accuracy (95% CI 35.0% to 63.0%; kappa = 0.233) versus 73.3% to 86.7% for LLMs (kappa = 0.600 to 0.800). Healthdirect operated under conservative interactive defaults while LLMs received complete information in a single prompt, which may have disadvantaged Healthdirect. Emergency sensitivity was 46.7% versus 80.0% to 86.7% for LLMs. Healthdirect produced two critical misses; no LLM produced any across 270 evaluations (95% CI 0% to 1.4%). When LLMs undertriaged, they recommended GP care rather than self-care. No tier differences were significant (all p > 0.05), and most systems over-triaged self-care cases.

**Interpretation:** Frontier LLMs demonstrated higher triage accuracy and safer error profiles than Healthdirect. All LLMs avoided critical misses; Healthdirect did not. Premium subscriptions did not significantly improve triage safety. These findings support clinical governance decisions about whether LLMs warrant formal evaluation alongside government-backed symptom checkers.

## 1. Introduction

Consumer-accessible large language models (LLMs) such as ChatGPT, Claude, and Gemini have become de facto advisors for clinical self-triage for millions of users worldwide. When a patient asks an AI chatbot whether chest pain warrants emergency care, the consequences of incorrect advice can be life-threatening. Under-triage, recommending self-care for an emergency, risks delayed treatment and death; over-triage, recommending emergency care unnecessarily, wastes resources but does not directly harm the patient. Ramaswamy et al. (2026) demonstrated in Nature Medicine that ChatGPT’s dedicated health interface undertriaged 52% of emergency cases,^1^ raising serious safety concerns. However, that study evaluated a single platform without comparison against existing tools, leaving a fundamental question unanswered: are LLMs safer or less safe than established services they are effectively displacing?

Government-backed symptom checkers represent the established alternative. Australia’s Healthdirect, a government-funded service recording 62 million community interactions in FY2024–25 including 2.4 million symptom checker uses, employs structured clinical algorithms under rigorous clinical governance.^2^ Yet accuracy has historically been modest: Semigran et al. (2015) found appropriate triage in only 57% of evaluations across 23 checkers,^3^ and Schmieding et al. (2022) found no improvement five years later.^4^ Hill et al. (2020) reported 49% accuracy across 36 Australian checkers including Healthdirect,^5^ though this predated consumer LLMs. Prior studies compared LLMs against NHS 111 Online^6^ and Ada and WebMD^7^ with mixed findings, but none directly evaluated Healthdirect against frontier LLMs.

A further gap concerns health equity. ChatGPT Plus and Claude Pro ($20/month) unlock more capable models unavailable on free tiers. If premium subscriptions offer meaningfully safer triage, AI-assisted clinical self-triage becomes dependent on ability to pay. The present study provides the first direct comparison of Healthdirect against six LLM configurations across three platforms and the first evaluation of whether premium subscriptions improve clinical self-triage safety.

## 2. Methods

### 2.1 Study design and ethics

This was a comparative, vignette-based evaluation study assessing the triage accuracy and safety of seven systems: Healthdirect and six LLM configurations across ChatGPT, Claude, and Gemini, each tested under free-tier and paid-tier conditions. The study used 45 standardised clinical vignettes from the Semigran et al. (2015) benchmark,^3^ adopted by Schmieding et al.^4^ in their five-year follow-up, enabling direct comparison with foundational studies in symptom checker evaluation.

This study used publicly available synthetic vignettes and did not involve human participants. No patient data were collected or used. Formal ethics approval was not required as the study involved no human subjects, identifiable data, or clinical intervention.

### 2.2 Clinical vignettes

Vignettes were originally compiled by Semigran et al.^3^ and expanded by Schmieding et al.^4^ We further adapted 25 of 45 vignettes to restore contextual information lost during abbreviation, including medication history, social history, and explicit negative symptoms. The remaining 20 were used as published. Adaptations did not alter clinical diagnosis or triage urgency; a subgroup comparison confirmed no significant difference in accuracy between adapted and unadapted vignettes across all seven systems (Fisher’s exact test, all p > 0.05; Supplementary Material E).

Each vignette was pre-assigned to one of three gold standard categories: emergency (n = 15; e.g., pulmonary embolism, myocardial infarction), non-emergent GP care (n = 15; e.g., urinary tract infection, otitis media), and self-care (n = 15; e.g., common cold, allergic rhinitis).^3^ All 45 vignettes were converted from third-person clinical language to first-person patient language following a standardised conversion rulebook (Supplementary Material B), replacing clinical terminology with everyday vocabulary and appending a fixed closing question: “What should I do — should I go to the emergency room, see a doctor, or can I handle this at home?”

### 2.3 Systems evaluated

Seven systems were evaluated across 315 total evaluations. Full model details and testing conditions are presented in Table 1. Model assignments were verified against each platform’s official documentation. For ChatGPT, the free-tier account does not display a model name to users; according to OpenAI’s Free Tier FAQ, free users access GPT-5.2.^8^ The Free Tier FAQ confirmed GPT-5.2 as the free tier model at the time of testing (accessed April 2026); subsequent changes to this page are consistent with OpenAI’s documented practice of updating model access on a rolling basis,^9^ as evidenced by the page’s active update timestamp (Supplementary Material H). The Plus account displayed GPT-5.3 Instant as the active model, confirmed via interface screenshot and official documentation^10^ (Supplementary Material G). For Claude, the free tier presents Sonnet 4.6, while Opus 4.7 is accessible only through a Pro subscription.^11^ For Gemini, both free and paid tiers provide access to the same models, with paid subscriptions offering higher usage limits rather than exclusive model access^12^; therefore, the Gemini comparison was framed as default versus manually selected best model rather than free versus paid. Model names and access conditions were recorded in April 2026 and may not reflect subsequent platform updates.

**Table 1.** Systems evaluated and testing conditions.

| System | Platform | Model | Selection Method | Access |
| --- | --- | --- | --- | --- |
| Healthdirect | healthdirect.gov.au | N/A | N/A | Free (public) |
| ChatGPT Free | chat.openai.com | GPT-5.2 (not displayed in UI) | Model assigned to free account | Free account |
| ChatGPT Plus | chat.openai.com | GPT-5.3 Instant (displayed in UI) | Model assigned to Plus account | Plus subscription (\$20/month) |
| Claude Free | claude.ai | Sonnet 4.6 | Model presented on free account | Free account |
| Claude Pro | claude.ai | Opus 4.7 | Manually selected (paid exclusive) | Pro subscription (\$20/month) |
| Gemini Default | gemini.google.com | Gemini 3 Flash | Model presented on free account | Free account |
| Gemini Pro | gemini.google.com | Gemini 3.1 Pro | Manually selected (available to all) | Free account |

### 2.4 Healthdirect evaluation protocol

Healthdirect was evaluated via its consumer web interface following a standardised seven-rule interaction protocol (Supplementary Material A), designed to ensure reproducible data entry. Key rules included entering only explicitly stated vignette information, defaulting to No for unmentioned symptoms on Yes/No questions, selecting the least severe and shortest duration options when not specified, and applying a three-tier symptom search strategy. Sessions were screen-recorded for verification. Healthdirect’s seven-level output was mapped to the three-tier classification as shown in Table 2. The mapping of “See a doctor within 2 hours” to EMERGENCY was tested in a sensitivity analysis with remapping to GP (Supplementary Material E).

**Table 2.** Healthdirect triage output mapping to three-tier classification.

| Healthdirect Output | Mapped To | Rationale |
| --- | --- | --- |
| Call 000 | EMERGENCY | Life-threatening emergency |
| Seek immediate medical care | EMERGENCY | Immediate urgency |
| See a doctor within 2 hours | EMERGENCY | Time-critical, requires urgent assessment |
| Call healthdirect | GP | Requires professional input |
| See a doctor within 24 hours | GP | Same-day or next-day GP |
| See a doctor within a week | GP | Routine GP visit |
| Self-care | SELF-CARE | Home management |

### 2.5 LLM evaluation protocol

All 270 LLM evaluations were conducted via each platform’s consumer web interface between April 17 and April 22, 2026. A fresh conversation was initiated for each evaluation, with memory and personalisation features disabled and separate accounts used for free-tier and paid-tier conditions. For ChatGPT Plus, Auto-switch to Thinking was disabled to ensure responses were generated by GPT-5.3 Instant rather than reasoning mode. Each evaluation consisted of a single first-person vignette prompt and one response with no follow-up questions or regeneration, reflecting typical user behaviour and consistent with prior comparable studies. All 270 sessions are preserved as publicly accessible shareable links (Supplementary Material D).

LLM responses were coded by a clinician co-author (B.C., formerly a practising physician) blinded to gold standard classifications, consistent with single-coder evaluation approaches used in prior comparable studies.^6,7^ Responses were randomised and stripped of case identifiers prior to coding. The primary recommendation was defined as the first actionable advice, excluding generic disclaimers. Conditional escalation advice, such as “see a doctor if symptoms worsen,” was not coded as the primary recommendation when the current advice was home management. Intra-coder reliability was assessed by re-coding 20 randomly selected responses after a two-week interval, yielding 100% agreement (kappa = 1.000). Model versions were recorded based on information displayed in each platform’s interface at the time of testing. For Gemini, the selected model is recorded in each shareable chat link. For Claude, the model displayed in the interface was noted at the start of each session.^11^ For ChatGPT, the Plus account displayed GPT-5.3 Instant; the free-tier model (GPT-5.2) was confirmed against OpenAI’s documentation as it is not displayed in the interface.^8,10^ Interface screenshots confirming model assignments are provided in Supplementary Material G.

### 2.6 Statistical analysis

Triage accuracy was defined as the proportion of vignettes matching the gold standard, with Wilson score 95% confidence intervals. Agreement was assessed using Cohen’s kappa.^13^ Cochran’s Q test assessed overall accuracy differences across systems. Pairwise comparisons used McNemar’s exact binomial test with Bonferroni correction (alpha = 0.0083). Safety metrics included emergency sensitivity, under-triage rate, critical miss rate, and over-triage rate. Critical miss rate was defined as the proportion of emergency cases triaged as self-care, representing the most dangerous error type. All analyses were performed in Python 3.

## 3. Results

### 3.1 Overall triage accuracy

Triage accuracy differed significantly across all seven systems (Cochran’s Q = 36.79, p < 0.001). Three LLM configurations significantly outperformed Healthdirect after Bonferroni correction (ChatGPT Free, p = 0.004; Claude Pro, p < 0.001; Gemini Pro, p < 0.001), while the remaining three showed significant differences at the uncorrected threshold but did not survive correction for multiple comparisons (Table 3). Healthdirect achieved 48.9% accuracy (kappa = 0.233), compared with 73.3% to 86.7% for LLMs (kappa = 0.600 to 0.800).

**Table 3.** Overall triage accuracy and agreement with gold standard.

| System | Correct/Total | Accuracy (%) | 95% CI | $\kappa$ | Agreement | McNemar p vs HD | Significance |
| --- | --- | --- | --- | --- | --- | --- | --- |
| Gemini Pro (3.1 Pro) | 39/45 | 86.7 | 73.8-93.7 | 0.800 | Almost perfect | <0.001 | Significant |
| Claude Pro (Opus 4.7) | 38/45 | 84.4 | 71.2-92.3 | 0.767 | Substantial | <0.001 | Significant |
| ChatGPT Free (GPT-5.2) | 35/45 | 77.8 | 63.7-87.5 | 0.667 | Substantial | 0.004 | Significant |
| Gemini Default (Flash) | 35/45 | 77.8 | 63.7-87.5 | 0.667 | Substantial | 0.011 | Not significant after correction |
| Claude Free (Sonnet 4.6) | 34/45 | 75.6 | 61.3-85.8 | 0.633 | Substantial | 0.017 | Not significant after correction |
| ChatGPT Plus (GPT-5.3) | 33/45 | 73.3 | 59.0-84.0 | 0.600 | Substantial | 0.013 | Not significant after correction |
| Healthdirect | 22/45 | 48.9 | 35.0-63.0 | 0.233 | Fair | — | — |

*Note: Bonferroni-corrected significance threshold: $\alpha=0.0083$ (0.05/6). 'Not significant after correction' indicates $p < 0.05$ but $p > 0.0083$ .*

**Table 4.** Safety metrics across all systems.

| System | Emergency Sensitivity | Under-Triage (n) | Critical Misses (n) | Over-Triage (n) |
| --- | --- | --- | --- | --- |
| Gemini Pro (3.1 Pro) | 86.7% (13/15) | 2 | 0 | 4 |
| Claude Pro (Opus 4.7) | 80.0% (12/15) | 3 | 0 | 4 |
| ChatGPT Free (GPT-5.2) | 80.0% (12/15) | 4 | 0 | 6 |
| Gemini Default (Flash) | 80.0% (12/15) | 3 | 0 | 7 |
| Claude Free (Sonnet 4.6) | 80.0% (12/15) | 3 | 0 | 8 |
| ChatGPT Plus (GPT-5.3) | 80.0% (12/15) | 4 | 0 | 8 |
| Healthdirect | 46.7% (7/15) | 13 | 2 | 10 |

### 3.2 Safety analysis

The most notable finding was the critical miss rate. Healthdirect produced two critical misses, whereas no LLM produced any across 270 evaluations (95% CI 0% to 1.4%). Healthdirect’s critical misses occurred in a 45-year-old man with acute flank pain consistent with renal colic (CASE_024) and a 65-year-old man with productive cough, fever, and comorbidities consistent with community-acquired pneumonia (CASE_026), both triaged as self-care. All six LLMs correctly identified CASE_024 as an emergency. For CASE_026, all LLMs recommended GP care rather than the gold standard of emergency, but substantially safer than Healthdirect’s self-care recommendation.

Healthdirect correctly identified 7 of 15 emergency cases (46.7%), compared with 12 to 13 of 15 for LLMs (80.0% to 86.7%). The three emergency cases most commonly missed by LLMs were asthma exacerbation unresponsive to inhaler (CASE_007), COPD exacerbation (CASE_022), and community-acquired pneumonia (CASE_026); in all instances, LLMs recommended GP care rather than self-care, representing a substantially less dangerous error.

### 3.3 Accuracy by triage category

Healthdirect showed lower accuracy across all three categories, achieving 46.7% on both emergency and GP cases and 53.3% on self-care cases. LLMs performed strongest on GP cases (86.7% to 100%) and emergency cases (80.0% to 86.7%), with the weakest performance on self-care cases (53.3% to 86.7%). Full category-level accuracy for all seven systems is presented in Supplementary Material F. All systems showed a tendency toward over-triage on self-care cases, consistent with prior findings in the symptom checker literature.^3,4^

### 3.4 Free versus paid model comparison

No statistically significant differences were observed between free-tier and paid-tier models on any platform (Table 5). Claude Pro and Gemini Pro showed approximately 9 percentage point improvements over their defaults, but neither reached significance (p = 0.219 and p = 0.125, respectively). A post hoc power analysis indicated that approximately 200 vignettes would be required to detect a 9-percentage-point difference at 80% power using McNemar’s test. These null results should therefore be interpreted as inconclusive rather than as evidence of no difference between tiers.

**Table 5.** Free-tier versus paid-tier model comparison.

| Platform | Free-tier Accuracy | Paid-tier Accuracy | Difference | McNemar p |
| --- | --- | --- | --- | --- |
| ChatGPT (GPT-5.2 vs GPT-5.3 Instant) | 77.8% | 73.3% | -4.4% | 0.625 |
| Claude (Sonnet 4.6 vs Opus 4.7) | 75.6% | 84.4% | +8.9% | 0.219 |
| Gemini (Flash vs 3.1 Pro) | 77.8% | 86.7% | +8.9% | 0.125 |

### 3.5 Cross-system agreement

In 14 cases, all six LLMs reached unanimous agreement on a triage recommendation that differed from Healthdirect. In 13 of these 14 cases (92.9%), the LLM recommendation matched the gold standard while Healthdirect’s did not. However, unanimous LLM agreement does not validate correctness, as it may reflect convergent biases or shared safety-oriented fine-tuning across models.

### 3.6 Sensitivity analysis

Remapping “See a doctor within 2 hours” from EMERGENCY to GP affected four cases. Healthdirect’s accuracy increased marginally from 48.9% to 53.3%, and the overall difference remained statistically significant (Cochran’s Q = 29.94, p < 0.001). Two LLM configurations remained significantly more accurate after Bonferroni correction. Critical miss findings were unaffected and primary conclusions unchanged (Supplementary Material E).

## 4. Discussion

### 4.1 Principal findings

This study provides the first direct comparison of Australia’s Healthdirect symptom checker against frontier LLMs for clinical self-triage safety. Three principal findings emerged. First, all six LLM configurations outperformed Healthdirect on triage accuracy, with three remaining significant after Bonferroni correction, and all LLMs avoided critical misses while Healthdirect produced two. When LLMs undertriaged emergency cases, they consistently recommended GP care rather than self-care, representing a meaningfully safer failure mode. Second, premium AI subscriptions did not significantly improve triage safety on any platform, suggesting that free-tier LLMs provide comparable clinical self-triage quality to paid alternatives. Third, all systems demonstrated a risk-averse tendency toward over-triage on self-care cases, consistent with prior symptom checker literature.^3,4^

### 4.2 Comparison with prior literature

Ramaswamy et al. (2026) reported that ChatGPT Health undertriaged 52% of emergency cases,^1^ considerably higher than the 20% observed here. This discrepancy likely reflects differences in vignette sets, prompt design, and the rapid evolution of underlying models. Ramaswamy used structured four-level output prompts across 960 assessments, whereas our study used naturalistic patient-language prompts without constraining the response format, which may elicit different triage behaviour. Fraser et al. (2023) concluded that unsupervised patient use of ChatGPT was not recommended, citing unsafe triage rates of 22%.^7^ Our results present a different picture three years later, with zero critical misses across 270 evaluations, though differences in vignette sets, prompt design, and comparator systems preclude direct attribution to model improvement alone. Healthdirect’s accuracy of 48.9% is consistent with Hill et al. (2020), who reported 49% across 36 Australian checkers,^5^ and falls within the range reported by Semigran et al.,^3^ suggesting Healthdirect’s performance is broadly representative of current symptom checker capabilities. Brown et al. (2025) similarly found comparable accuracy between LLMs and NHS 111 Online with LLMs demonstrating a safer error direction,^6^ and Kopka et al. (2026) reported substantial variability across ChatGPT model versions,^14^ consistent with the cross-platform patterns observed here.

### 4.3 Methodological asymmetry

A critical distinction must be acknowledged between the two evaluation formats. Healthdirect was tested interactively under conservative protocol defaults, with unanswered questions defaulting to No, severity to least severe, and duration to shortest. These defaults systematically constrained the information available to its algorithm. LLMs received complete clinical information in a single prompt. The higher LLM accuracy therefore reflects performance under relatively complete information conditions, which may have disadvantaged Healthdirect. In real-world settings, Healthdirect’s structured questioning would elicit genuine patient responses rather than conservative defaults, potentially yielding better performance. Conversely, real patients may describe symptoms less coherently than the structured vignettes used here, potentially reducing LLM accuracy. Both directions of this asymmetry should be considered when interpreting the performance gap. The LLM-specific direction of this asymmetry is discussed further in Section 4.7.

### 4.4 Clinical and policy implications

These findings have direct implications for clinical governance and digital health policy. The substantial performance gap and the critical miss finding provide evidence to support formal evaluation of whether LLMs should be integrated alongside or as complements to government-backed symptom checker services. Health authorities considering such integration should prioritise prospective clinical studies with real patient interactions to validate these vignette-based findings before deployment at scale.

The absence of a statistically significant free-versus-paid safety advantage is reassuring from an equity perspective, but should be interpreted cautiously given the underpowered comparison. Consumers who cannot afford premium AI subscriptions do not appear to receive meaningfully inferior triage advice, at least in the current generation of models. However, the underpowered free-versus-paid comparison means this finding should be interpreted as inconclusive pending larger studies.

The universal over-triage tendency on self-care cases has system-level implications. If LLMs are adopted at scale as triage advisors, even a modest increase in unnecessary GP presentations across a population of millions of users could place meaningful additional demand on primary care services. This risk warrants consideration in any governance framework for consumer AI in health.

### 4.5 Failure mode analysis

The qualitative nature of errors differed substantially between systems. Healthdirect produced multidirectional errors, both over-triaging conditions such as strep pharyngitis and under-triaging emergencies such as renal colic and community-acquired pneumonia. LLM errors were predominantly unidirectional, overwhelmingly over-triaging self-care conditions as requiring GP consultation. This risk-averse pattern likely reflects safety-oriented fine-tuning applied across all major LLMs. While unidirectional over-triage is a safer failure profile than Healthdirect’s bidirectional errors, it is not without consequence at population scale.

### 4.6 Methodological strengths

This study offers several methodological contributions. Blinded coding with randomised response ordering eliminated confirmation bias in triage classification, with intra-coder reliability confirmed at kappa = 1.000 on a random 20-response subsample. All 270 LLM sessions are publicly accessible via shareable links, providing a level of transparency rarely seen in this field and enabling independent verification. The seven-rule Healthdirect interaction protocol provides a reproducible standard for future symptom checker evaluations.

### 4.7 Limitations

Several limitations should be considered. First, vignette-based evaluation disproportionately constrains interactive systems such as Healthdirect, whose real-world performance with actual patients may exceed the results reported here. Second, the 45-vignette sample, while an established benchmark,^3,4^ limited statistical power for the free-versus-paid comparisons. Third, LLMs evolve rapidly and these results represent a snapshot of April 2026 capabilities; future model versions may perform differently. Single-session evaluation means stochastic variation across runs is unaccounted for, though this approach is consistent with prior studies.^6,7^ Fourth, this study assessed triage recommendations in isolation and did not evaluate the quality of accompanying clinical explanations or differential diagnoses. Fifth, single-prompt evaluation may not reflect real patient interactions, where symptoms are described incrementally and ambiguously across multiple messages. Multi-turn conversational evaluation represents an important direction for future research. Prospective studies evaluating LLM triage safety in real clinical settings, including emergency department and primary care contexts, would provide the real-world evidence needed to inform deployment decisions.

## 5. Conclusions

In this standardised vignette evaluation, frontier LLMs (ChatGPT, Claude, and Gemini) demonstrated higher triage accuracy and safer error profiles than Healthdirect when LLMs received complete symptom descriptions and Healthdirect was tested interactively under conservative protocol defaults. The most critical safety finding was that all LLMs avoided the most dangerous error type, triaging emergency patients as self-care, while Healthdirect did not. No statistically significant improvement in triage safety was observed between free-tier and paid-tier models on any platform, although the sample size may have been insufficient to detect the observed differences.

These findings indicate that the current generation of consumer-accessible LLMs can produce fewer critical safety failures than a government-backed symptom checker under the conditions evaluated. However, neither LLMs nor symptom checkers achieved perfect accuracy, and all systems showed a tendency toward risk-averse over-triage. These results should not be interpreted as endorsing unsupervised clinical use of LLMs, but rather as evidence supporting formal prospective evaluation of LLMs in real clinical settings before governance decisions are made about their role in consumer health services.

## Data Availability

All data produced in the present study are contained in the manuscript and its supplementary materials (Supplementary Materials A-H).

## CRediT authorship contribution statement

**Azwad Raza Chowdhury:** Conceptualization, Data curation, Formal analysis, Investigation, Methodology, Project administration, Visualization, Writing – original draft, Writing – review and editing. **Bushra Chowdhury:** Investigation, Methodology, Validation, Writing – review and editing.

## Ethics approval and consent to participate

Formal ethics approval was not required. The research was conducted using publicly available synthetic clinical vignettes and did not involve human participants, patient data, biological materials, or clinical intervention. No personally identifiable information was collected or used at any stage of the study.

## Declaration of generative AI and AI-assisted technologies in the manuscript preparation process

During the preparation of this work, the authors used Claude (Anthropic) to assist with manuscript drafting, editing, and structural organisation. After using this tool, the authors reviewed and edited the content as needed and take full responsibility for the content of the manuscript.

## Data availability

All LLM evaluation sessions are publicly accessible via shareable links provided in Supplementary Material D. Adapted clinical vignettes and the conversion rulebook are available in Supplementary Material B. The Healthdirect interaction protocol is provided in Supplementary Material A. The response coding guide is provided in Supplementary Material C. Subgroup and sensitivity analyses are provided in Supplementary Material E. Category-level accuracy data for all seven systems are provided in Supplementary Material F. Interface screenshots confirming model assignments during testing are provided in Supplementary Materials G and H. No patient data were collected or used in this study.

## Acknowledgements

The authors thank Emily Danvers for her guidance on research design and manuscript preparation.

