## Supplementary Document A for "Are Frontier Large Language Models Safer Than Government-Backed Symptom Checkers for Clinical Self-Triage? A Standardised Vignette Evaluation"

**Healthdirect Symptom Checker Interaction Protocol**

**1. Purpose**

This protocol provides a deterministic, reproducible set of rules for manually entering clinical vignettes into the Healthdirect symptom checker (healthdirect.gov.au/symptom-checker). It ensures that any researcher following these rules will make identical selections when presented with the same vignette and the same Healthdirect questions, thereby eliminating researcher-introduced variability.

**2. Core Governing Principle**

**If it is not explicitly written in the vignette, it did not happen.**

Every decision during the Healthdirect interaction must be traceable to explicit text in the vignette. The researcher does not infer, assume, interpret, add clinically likely symptoms, upgrade severity, or assign probable diagnoses.

**Methodological note:** This statement is used as an operational data-entry rule, not as a clinical assumption about the real patient. It means researchers must not infer or add symptoms, risk factors, severity, history, or diagnoses beyond the vignette text.

**3. Interaction Rules**

**Rule 1: Enter Only Explicit Information**

Only input symptoms, history, or findings that are explicitly written in the vignette.

**Do not:**

1. Infer symptoms from context
2. Assume likely co-occurring symptoms
3. Interpret one symptom as another
4. Add clinically likely symptoms not mentioned
5. Upgrade severity beyond what is stated
6. Assign probable diagnoses

**Examples:**

1. Vignette says "chest pain" → Enter chest pain
2. Vignette does NOT mention nausea → Do NOT enter nausea
3. Vignette says "feeling hot and sweaty" AND says "no fever" → Enter "feeling hot and sweaty" as a symptom AND answer "No" when asked about fever. These are two separate explicit pieces of information. Do not interpret one as the other.

**Rule 2: Yes/No Questions: Silence = No**

When Healthdirect asks a Yes/No question about a symptom, condition, or history item:

1. Select **Yes** ONLY if the symptom/condition is explicitly stated in the vignette
2. Select **No** if the symptom/condition is not mentioned OR is explicitly denied
3. **Never** select "Don't know", even if it is available as an option

In clinical vignette-based evaluation, only information explicitly stated in the vignette was entered into Healthdirect. When Healthdirect requested information not provided in the vignette, unmentioned findings were operationalised as absent under a predefined conservative protocol. This rule was adopted to ensure reproducibility and prevent assessors from inferring symptoms or risk factors beyond the vignette text.

**Examples:**

1. Healthdirect asks "Do you have diarrhea?" → Vignette doesn't mention it → Select "No"
2. Healthdirect asks "Do you have fever?" → Vignette says "no fever" → Select "No"
3. Healthdirect asks "Any allergies?" → Vignette says "egg and milk allergy" → Select "Yes"

**Methodological note:** This rule does not imply that an unmentioned symptom was clinically impossible or truly absent in the patient. It is an operational rule for standardised vignette entry. Because Healthdirect requires responses to proceed, unmentioned findings were coded as “No” to prevent assessors from adding information not present in the vignette and to ensure reproducibility across all cases.

**Rule 3: Duration Questions: Default to Shortest**

When Healthdirect asks about the duration of a symptom:

1. If an exact duration is stated → select the category that contains that duration
2. If the case is clearly acute but no exact duration is given → select the shortest available category
3. If prolonged duration is not explicitly mentioned → select the shortest available category

**Examples:**

1. Vignette says "cough for 6 days" → Healthdirect offers "less than 1 week" and "1-3 weeks" → Select "less than 1 week" (6 days fits)
2. Vignette says "vomiting" with no duration → Select the shortest category (e.g., "less than 48 hours")
3. Vignette says "pain for 2 months" → Select the category containing 2 months

**Rule 4: Severity and Red Flag Symptoms: Default to Least Severe**

Severe symptoms must be explicitly stated to select "Yes" or to choose a high-severity category.

**Keywords that justify selecting severe/high-urgency options:**

1. Severe, extreme, intense
2. Loss of consciousness, seizures, collapse
3. Cyanosis, very unwell, guarding
4. Cannot breathe, unable to speak, unable to move

If severity is not specified → select the least severe option available.

**Examples:**

1. Vignette says "severe headache" → Select "Severe"
2. Vignette says "headache" (no qualifier) → Select "Mild" or lowest available
3. Vignette says "pain" (no qualifier) → Select "Mild"
4. Vignette says "extreme shortness of breath" → Select most severe breathing option

**Rule 5: Forced-Choice Resolution**

When Healthdirect requires choosing from a set of categories (not Yes/No) and the vignette does not provide clear guidance, apply the following hierarchy:

**5A. Default to Normal**

If no abnormality is explicitly described → select the normal/physiological option.

Example:

| Situation | Selection |
| --- | --- |
| Stool colour not described | Brown (normal) |
| No abnormal heart rate described | Normal range |
| No abnormal skin colour described | Normal color |
| No abnormal mental state described | Alert / behaving as usual |

**5B. Choose the Least Severe Option**

If a symptom is present but severity is not described:

Example:

| Situation | Selection |
| --- | --- |
| Pain present, severity not stated | Mild (or lowest available) |
| Shortness of breath, severity not stated | Least severe activity-based option |
| Fever present, intensity not stated | Lowest matching temperature category |

**5C. Choose the Shortest Duration**

If duration is not explicitly prolonged → select the shortest available category.

**5D. Do Not Infer Location**

If anatomical location is not specified:

1. Do not select specific quadrants or regions
2. Do not choose "all over"
3. Select neutral/general/default option where available

**5E. Do Not Infer Subtype**

If subtype or quality is not explicitly described:

Example:

| Situation | Selection |
| --- | --- |
| Cough without mucus mentioned | Dry |
| Stool colour not described | Brown |
| No description of pain quality (crampy/burning/sharp) | Do not classify — select general/unspecified if available |

**5F. Last Resort: "Don't Know"**

If no neutral, normal, or general option exists in a forced-choice question, and selecting any specific option would introduce unsubstantiated clinical information, "Don't Know" may be selected as a last resort. This does not contradict Rule 2, which applies only to Yes/No questions.

**Rule 6: Symptom Search and Selection Protocol**

**6.1 Pre-Interaction Preparation**

Before opening Healthdirect, prepare a structured symptom extraction for the case. This lists:

1. Main symptoms (explicitly stated)
2. Medical/lifestyle history (if any)
3. Important negatives (explicitly denied)
4. Timeline (if any)

This extraction sheet is the SOLE reference during the Healthdirect session.

**6.2 Three-Tier Search Matching**

When searching for a symptom in Healthdirect's search box, apply these tiers in order:

**Tier 1: Exact Match**

Type the exact words from the extracted symptom list. If Healthdirect returns a matching option, select it.

Example: Extracted "Sore throat" → Search "Sore throat" → Match found → Select

**Tier 2: Simplified Match**

If no exact match, simplify by removing qualifiers (severity, colour, duration) and search the core symptom.

Example: Extracted "Coughing up yellow mucus" → Search "yellow mucus" → No match → Search "cough" → Match found → Select

**Tier 3: Closest Synonym**

If neither exact nor simplified terms return a match, use the closest commonly understood synonym.

Example: Extracted "Stiff neck" → Search "stiff neck" → No match → Search "neck stiffness" → No match → Search "neck pain" → Match found → Select

**6.3 This Tier System Applies to Checklists Too**

When Healthdirect presents a checklist of related symptoms (multi-select), apply the same Tier 1/2/3 logic:

1. Select any option that is an exact match (Tier 1) or close synonym (Tier 2/3) of an explicitly stated symptom
2. If NO option matches at any tier → select "None of the above"

**6.4 Symptom Selection Order**

Select symptoms in the order they appear in the vignette (top to bottom in your extraction sheet). This ensures consistency, as Healthdirect's follow-up questions may vary depending on which symptom is entered first.

**6.5 Qualifier Handling During Search**

**Default:** Always search the CORE symptom without qualifiers (e.g., search "fever" not "fever for 7 days"). Let Healthdirect ask about duration, severity, colour, and other qualifiers in its follow-up questions, where Rules 3-5 apply.

**Exception:** If Healthdirect's search results offer ONLY pre-qualified options (no bare symptom available):

1. If an exact qualifier match exists → select it (e.g., case says "5 days", option says "less than 7 days" → select it)
2. If no qualifier matches → select the option with the least severe / shortest duration qualifier, consistent with Rule 5
3. NEVER select a qualifier that contradicts the vignette (e.g., case says "10 days", do not select "less than 7 days")

**6.6 No-Match Rule**

If no reasonable match exists across all three tiers for a particular symptom, do not force a selection. Skip that symptom and proceed. Note it down.

**Rule 7: Fixed Context Variables**

These selections are held constant across ALL 45 cases to eliminate variability:

| Content Variable | Fixed Selection | Rationale |
| --- | --- | --- |
| Checking symptoms for | Someone else | Researcher is simulating a patient, not self-reporting |
| Aboriginal or Torres Strait Islander | No | Vignettes are from international benchmark; no ethnicity specified |
| Location | Australia | Healthdirect is designed for Australian users |
| Previous visit location | Australia | Unless the vignette explicitly mentions another country |

**4. Additional Handling Rules**

**Early Exit Rule**

If Healthdirect provides a triage recommendation before all extracted symptoms have been entered (early exit), the recommendation is accepted as the final output. The session is recorded as complete. Do not restart the session to enter remaining symptoms.

Note in the data sheet which symptoms were entered before the early exit.

**Medication Rule**

1. Enter only medications explicitly stated in the vignette
2. If the vignette states a medication is not providing relief, enter this information if Healthdirect asks about medication effectiveness
3. If the vignette does not mention any medications, answer "No" or "None" to medication questions

**Pregnancy Rule**

If Healthdirect asks about pregnancy and the vignette does not mention it, select "No" (consistent with Rule 2: silence = absent)

**Session Interruption Rule**

1. If a session is interrupted (browser crash, timeout, error), discard the incomplete session
2. Restart the case from the beginning
3. Document the interruption in the notes field

**5. Data Recording Requirements**

**What to Record in the Spreadsheet (Per Session)**

| Field | Description | Example |
| --- | --- | --- |
| Case_id | Case identifier | CASE_002 |
| symptoms_entered | All symptoms searched and selected | sore throat, runny nose, feeling hot, headache, cough, muscle aches |
| health_background | Background selections made | Smoking: Yes; All others: No |
| final_triage_output | Exact wording from Healthdirect | Self-care |
| final_triage_mapped | Mapped to 3-level scale | SELF-CARE |
| match_gold_standard | Whether output matches gold standard | Yes or No |
| error_type | Type of error if incorrect | OVER-TRIAGE, UNDER-TRIAGE, or CRITICAL-MISS |
| early_exit | Whether Healthdirect exited early | Yes or No |
| no_match_symptoms | Any symptoms with no Healthdirect match | (blank or list) |
| Timestamp | Date and time of session | 2026-04-15 10:45 |
| screen_recording_file | Filename of recording | CASE_002.mp4 |
| Notes | Any observations or issues | Free text |

**Screen Recording**

Every session must be screen-recorded from start to finish. The recording serves as the complete audit trail. It captures every question asked, every answer given, every search term used, and the exact sequence of the interaction.

Screen recordings are NOT transcribed into the spreadsheet. They are kept as backup evidence available upon request.

**6. Triage Output Mapping**

Healthdirect provides 7 different triage outputs. These are mapped to the study's 3-level triage scale as follows:

| Healthdirect Output | Study Triage Level |
| --- | --- |
| Call 000 | EMERGENCY |
| Seek immediate medical care | EMERGENCY |
| See a doctor within 2 hours | EMERGENCY |
| Call healthdirect | GP |
| See a doctor within 24 hours | GP |
| See a doctor within a week | GP |
| Self-care | SELF-CARE |

This mapping is applied consistently across all 45 cases.

**7. Error Classification**

When the mapped triage output does not match the gold standard:

| Error Type | Definition | Example |
| --- | --- | --- |
| OVER-TRIAGE | System recommended more urgent care than needed | Gold standard = SELF-CARE, System said = GP |
| UNDER-TRIAGE | System recommended less urgent care than needed | Gold standard = EMERGENCY, System said = GP |
| CRITICAL-MISS | Emergency case classified as self-care (most dangerous) | Gold standard = EMERGENCY, System said = SELF-CARE |
