## Supplementary Document E for "Are Frontier Large Language Models Safer Than Government-Backed Symptom Checkers for Clinical Self-Triage? A Standardised Vignette Evaluation"

**Supplementary Material E: Subgroup and Sensitivity Analyses**

**1. Subgroup Analysis: Adapted Versus Unadapted Vignettes**

**Purpose**

To determine whether the adaptation of 25 vignettes (restoring patient-reportable contextual information) introduced systematic bias in vignette difficulty compared with the 20 unadapted vignettes used as published by Schmieding et al.

**Method**

The 45 vignettes were classified into two groups: adapted (n = 25), where contextual information such as medication history, social history, family history, and explicit negative symptoms were restored; and unadapted (n = 20), which were used as published by Schmieding et al. without modification. Triage accuracy was calculated separately for each group across all seven systems. Fisher's exact test was used to compare accuracy between groups for each system.

**Gold Standard Distribution**

| Triage Category | Adapted (n = 25) | Unadapted (n = 20) |
| --- | --- | --- |
| Emergency | 9 | 6 |
| GP | 8 | 7 |
| Self-care | 8 | 7 |

**Results**

**Table E1**. Triage accuracy for adapted versus unadapted vignettes by system.

| System | Adapted (n = 25) | Unadapted (n = 20) | Difference | Fisher’s p |
| --- | --- | --- | --- | --- |
| Healthdirect | 14/25 (56.0%) | 8/20 (40.0%) | +16.0pp | 0.373 |
| ChatGPT Free (GPT-5.2) | 20/25 (80.0%) | 15/20 (75.0%) | +5.0pp | 0.731 |
| ChatGPT Plus (GPT-5.3) | 19/25 (76.0%) | 14/20 (70.0%) | +6.0pp | 0.741 |
| Claude Free (Sonnet 4.6) | 20/25 (80.0%) | 14/20 (70.0%) | +10.0pp | 0.500 |
| Claude Pro (Opus 4.7) | 22/25 (88.0%) | 16/20 (80.0%) | +8.0pp | 0.682 |
| Gemini Default (Flash) | 21/25 (84.0%) | 14/20 (70.0%) | +14.0pp | 0.301 |
| Gemini Pro (3.1 Pro) | 23/25 (92.0%) | 16/20 (80.0%) | +12.0pp | 0.383 |

Overall accuracy: Adapted 79.4% (139/175) vs Unadapted 69.3% (97/140).

**Conclusion**

No statistically significant difference in triage accuracy was observed between adapted and unadapted vignettes for any of the seven systems (Fisher's exact test, all p>0.05). These findings did not provide evidence that restoration of contextual information introduced systematic bias in vignette difficulty.

**2. Sensitivity Analysis: Alternative Triage Mapping for "See a Doctor Within 2 Hours"**

**Purpose**

To assess whether mapping Healthdirect's "See a doctor within 2 hours" output to EMERGENCY (rather than GP) unduly influenced the primary conclusions of the study.

**Method**

In the primary analysis, "See a doctor within 2 hours" was mapped to EMERGENCY. In this sensitivity analysis, this output level was remapped to GP, and all accuracy metrics, statistical tests, and safety metrics were recalculated. LLM results were unaffected by this remapping as it applies only to Healthdirect's output categories

**Cases Affected**

Four Healthdirect evaluations produced a "See a doctor within 2 hours" output:

**Table E2**. Cases affected by the alternative triage mapping.

| Case | Gold Standard | Primary Mapping (EMERGENCY) | Alternative Mapping (GP) | Effect on Accuracy |
| --- | --- | --- | --- | --- |
| CASE_008 | EMERGENCY | Correct | Incorrect | -1 |
| CASE_032 | GP | Incorrect (over-triage) | Correct | +1 |
| CASE_036 | GP | Incorrect (over-triage) | Correct | +1 |
| CASE_041 | GP | Incorrect (over-triage) | Correct | +1 |

Net effect: Healthdirect gains 2 correct classifications (3 gained minus 1 lost).

**Results: Accuracy Comparison**

**Table E3**. Healthdirect accuracy under primary versus alternative mapping.

| Metric | Primary Mapping | Alternative Mapping | Change |
| --- | --- | --- | --- |
| Overall accuracy | 22/45 (48.9%) | 24/45 (53.3%) | +4.4pp |
| Emergency sensitivity | 7/15 (46.7%) | 6/15 (40.0%) | -6.7pp |
| Under-triage | 13 | 14 | +1 |
| Critical misses | 2 | 2 | Unchanged |
| Over-triage | 10 | 7 | -3 |
| Cohen's kappa | 0.233 | 0.300 | +0.067 |

**Results: Statistical Tests**

**Table E4.** Comparison of statistical significance under primary versus alternative mapping.

| Test | Primary Mapping | Alternative Mapping | Conclusion Changed? |
| --- | --- | --- | --- |
| Cochran's Q (overall) | Q=36.79, p<0.001 | Q=29.94, p<0.001 | No |
| ChatGPT Free vs HD | p=0.004 (Sig) | p=0.013 (Not sig after Bonferroni) | Minor change |
| ChatGPT Plus vs HD | p=0.013 (Not sig after Bonferroni) | p=0.035 (Not sig after Bonferroni) | No |
| Claude Free vs HD | p=0.017 (Not sig after Bonferroni) | p=0.041 (Not sig after Bonferroni) | No |
| Claude Pro vs HD | p<0.001 (Sig) | p=0.001 (Sig) | No |
| Gemini Default vs HD | p=0.011 (Not sig after Bonferroni) | p=0.027 (Not sig after Bonferroni) | No |
| Gemini Pro vs HD | p<0.001 (Sig) | p<0.001 (Sig) | No |

Under the alternative mapping, two LLM configurations (Claude Pro, Gemini Pro) remained significantly more accurate than Healthdirect after Bonferroni correction, compared with three (ChatGPT Free, Claude Pro, Gemini Pro) under the primary mapping.

**Results: Safety Metrics**

The critical miss count remained unchanged at 2 under both mappings (CASE_024 and CASE_026). The finding of zero LLM critical misses across 270 evaluations was unaffected by the remapping, as the sensitivity analysis applies only to Healthdirect's output categories.

**Conclusion**

The alternative triage mapping resulted in a marginal improvement in Healthdirect's accuracy (+4.4pp) but did not alter the primary conclusions of the study. The overall difference across systems remained highly significant (p<0.001), LLMs continued to demonstrate higher accuracy and safer error profiles than Healthdirect, and the critical safety findings (zero LLM critical misses versus two Healthdirect critical misses) were unchanged.
