## Supplementary Document F for "Are Frontier Large Language Models Safer Than Government-Backed Symptom Checkers for Clinical Self-Triage? A Standardised Vignette Evaluation"

**Supplementary Material F: Accuracy by Triage Category**

This supplementary material presents category-level triage accuracy for all seven systems across the three gold standard categories: emergency, non-emergent GP care, and self-care. Overall accuracy is included for reference. These data are referenced in Section 3.3 of the main manuscript.

**Table F1**. Triage accuracy by category for all seven systems.

| System | Emergency (n = 15) | GP (n = 15) | Self-care (n = 15) | Overall Accuracy |
| --- | --- | --- | --- | --- |
| Gemini Pro (3.1 Pro) | 86.7% | 100.0% | 73.3% | 86.7% |
| Claude Pro (Opus 4.7) | 80.0% | 86.7% | 86.7% | 84.4% |
| ChatGPT Free (GPT-5.2) | 80.0% | 86.7% | 66.7% | 77.8% |
| Gemini Default (Flash) | 80.0% | 93.3% | 60.0% | 77.8% |
| Claude Free (Sonnet 4.6) | 80.0% | 93.3% | 53.3% | 75.6% |
| ChatGPT Plus (GPT-5.3) | 80.0% | 86.7% | 53.3% | 73.3% |
| Healthdirect | 46.7% | 46.7% | 53.3% | 48.9% |

*Note: Accuracy calculated as the proportion of vignettes correctly classified within each category (n = 15 per category). Overall accuracy calculated across all 45 vignettes. Systems ordered by overall accuracy (highest to lowest).*
