## Supplementary Document G for "Are Frontier Large Language Models Safer Than Government-Backed Symptom Checkers for Clinical Self-Triage? A Standardised Vignette Evaluation"

**Supplementary Material G: ChatGPT Plus Model Assignment Screenshot**

This supplementary material provides supporting evidence that GPT-5.3 Instant was the active model assigned to the ChatGPT Plus account used during evaluation. The screenshot was captured after the study testing period during personal use; therefore, it is provided to document model assignment rather than the exact evaluation settings. Official OpenAI documentation also identifies GPT-5.3 Instant as a model available to paid ChatGPT users during this period [10].

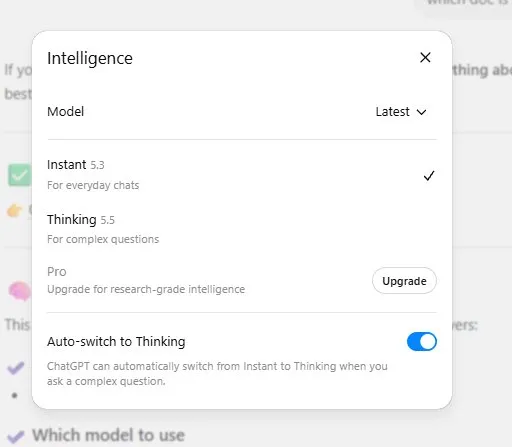

Figure G1. Screenshot of the ChatGPT Plus model picker interface. The interface shows GPT-5.3 Instant selected as the active model. The screenshot was captured after the study testing period and shows the Auto-switch to Thinking toggle enabled. During all study evaluations, this toggle was disabled to ensure responses were generated by GPT-5.3 Instant without automatic escalation to reasoning mode. The screenshot is provided as interface-level evidence of model assignment, while Reference [10] provides official documentation of GPT-5.3 Instant availability.
