## Supplementary Document H for "Are Frontier Large Language Models Safer Than Government-Backed Symptom Checkers for Clinical Self-Triage? A Standardised Vignette Evaluation"

**Supplementary Material H: Evidence for ChatGPT Free Tier Model Assignment (GPT-5.2) During Testing Period**

This supplementary material provides documentary evidence supporting the classification of GPT-5.2 as the active ChatGPT free-tier model during the study testing window of April 17–22, 2026. It addresses the inherent limitation that OpenAI's help pages are updated on a rolling basis and may not retain historical content at the time of later access.

**H1. ChatGPT Free Tier FAQ supporting GPT-5.2 (captured April 29, 2026)**

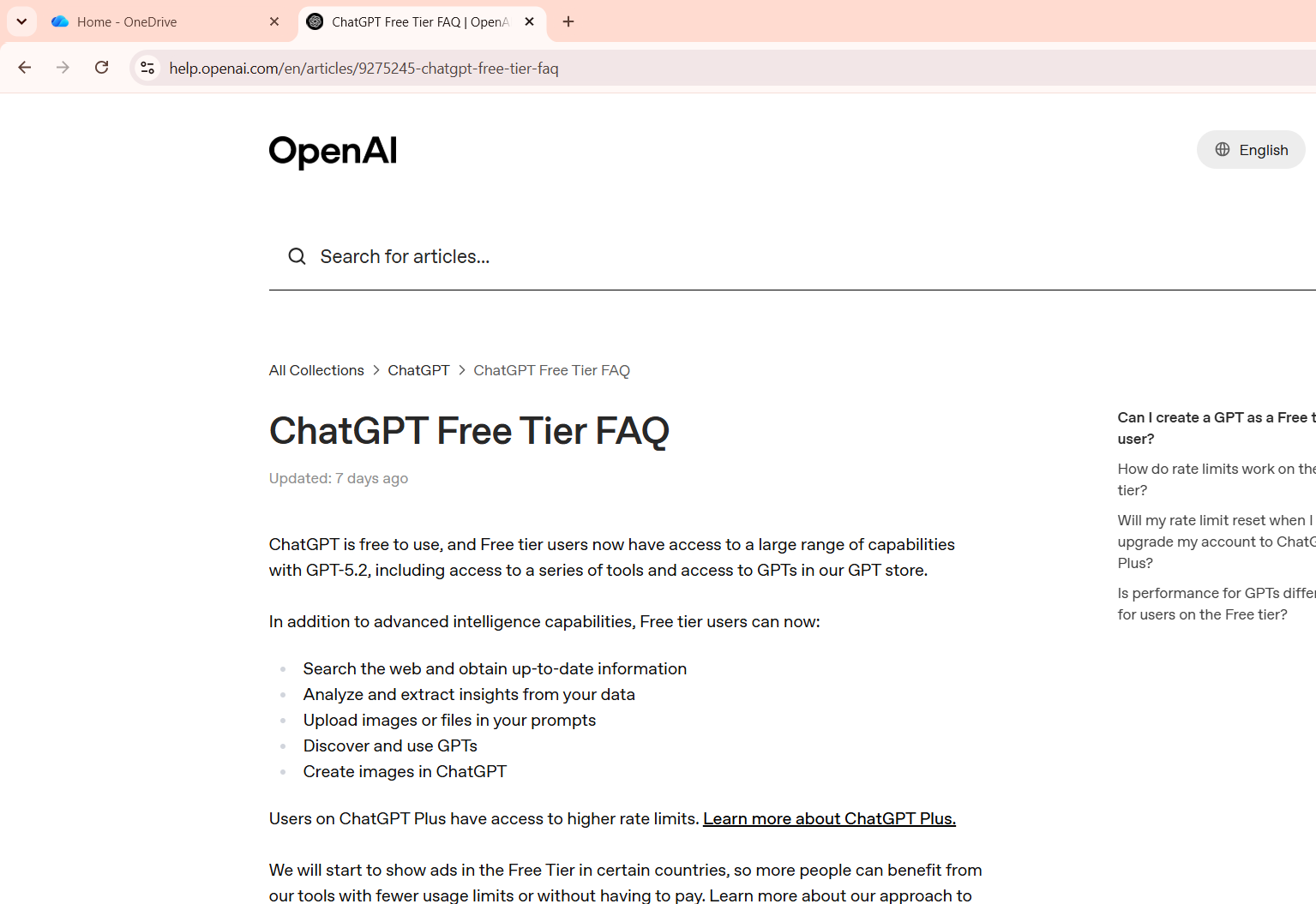
Figure H1. Screenshot of the OpenAI ChatGPT Free Tier FAQ page (help.openai.com/en/articles/9275245-chatgpt-free-tier-faq) captured on April 29, 2026, seven days after the testing window closed. The page explicitly states GPT-5.2 as the free-tier model. The "Updated: 7 days ago" timestamp indicates the page was last updated approximately April 22, 2026, which falls within the testing window of April 17-22, 2026. The full URL is visible in the browser address bar confirming the source page.

**H2. Confirmation that the Free Tier FAQ page is updated frequently (captured June 6, 2026)**

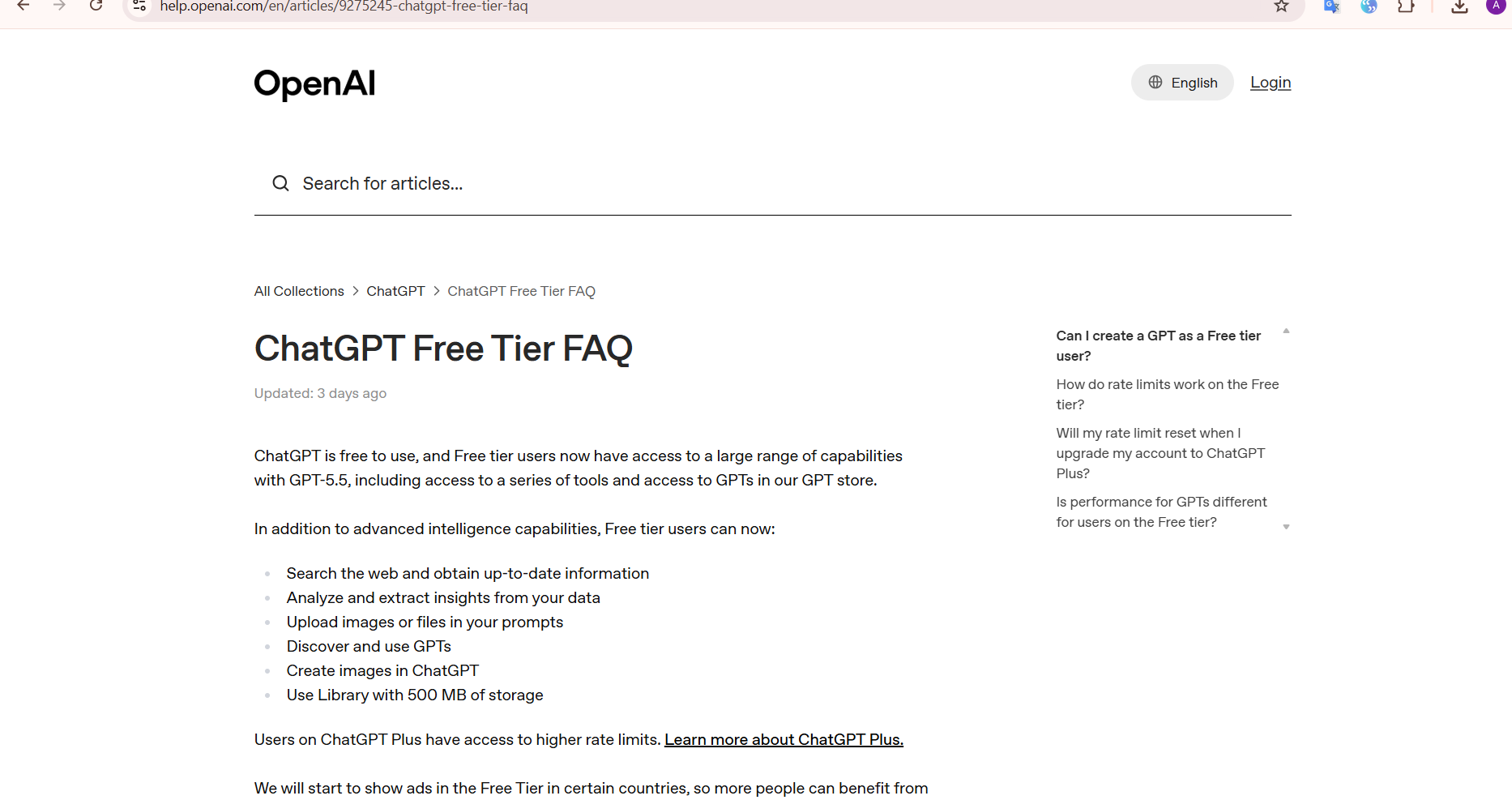
Figure H2. Screenshot of the same ChatGPT Free Tier FAQ page captured on a later date, showing "Updated: yesterday." This supports that OpenAI updates this page on a rolling basis as models change [9], explaining why the current page content no longer reflects the GPT-5.2 information visible in Figure H1. This pattern of rolling updates is consistent with the access date recorded in the main manuscript reference [8].

**H3. Model release timeline supporting GPT-5.2 was active during testing**

The following timeline was compiled from official OpenAI documentation and contemporaneous screenshots captured during and shortly after the testing period, and supports the conclusion that GPT-5.2 was the active free-tier ChatGPT model throughout the testing window of April 17-22, 2026.

| Date | Event | Implication for study |
| --- | --- | --- |
| December 11, 2025 | GPT-5.2 released; release notes state that free users use GPT-5.2 Instant by default [9] | Supports GPT-5.2 as the default free-tier model before the April 2026 testing window |
| March 5, 2026 | GPT-5.3 Instant released for Plus and paid users only [10] | Free tier remains on GPT-5.2 |
| April 17-22, 2026 | Study testing window | GPT-5.2 active free-tier model per FAQ (Figure H1) |
| April 29, 2026 | Free Tier FAQ screenshot captured (Figure H1) | Page stating GPT-5.2, updated approximately April 22 per timestamp |

Taken together, the FAQ screenshot (Figure H1), the rolling update evidence (Figure H2), and the model release timeline (Section H3) support the conclusion that GPT-5.2 was the active free-tier ChatGPT model during the study testing window. The subsequent disappearance of this information from the live FAQ page is consistent with OpenAI's documented practice of updating help pages as models change.
